# D614G Spike Mutation Increases SARS CoV-2 Susceptibility to Neutralization

**DOI:** 10.1101/2020.07.22.20159905

**Authors:** Drew Weissman, Mohamad-Gabriel Alameh, Thushan de Silva, Paul Collini, Hailey Hornsby, Rebecca Brown, Celia C. LaBranche, Robert J Edwards, Laura Sutherland, Sampa Santra, Katayoun Mansouri, Sophie Gobeil, Charlene McDanal, Norbert Pardi, Nick Hengartner, Paulo J.C. Lin, Ying Tam, Pamela A. Shaw, Mark G. Lewis, Carsten Boesler, Uğur Şahin, Priyamvada Acharya, Barton F. Haynes, Bette Korber, David C. Montefiori

**Affiliations:** Division of Infectious Diseases, University of Pennsylvania Perelman School of Medicine, Philadelphia, PA USA; Department of Infection, Immunity and Cardiovascular Disease, University of Sheffield, Sheffield, UK; South Yorkshire Regional Department of Infection and Tropical Medicine, Sheffield Teaching Hospitals NHS Foundation Trust, Sheffield, UK; Department of Surgery, Duke University School of Medicine, Durham, North Carolina USA; Duke Human Vaccine Institute, Duke University School of Medicine, Durham, NC USA; Duke University, Department of Medicine, Durham NC USA; Center for Virology and Vaccine Research, Beth Israel Deaconess Medical Center, Harvard Medical School, Boston, Massachusetts USA; T6: Theoretical Biology and Biophysics, Los Alamos National Laboratory, Los Alamos, NM USA; Acuitas Therapeutics, Vancouver, BC, CA; Department of Biostatistics, Epidemiology and Informatics University of Pennsylvania Perelman School of Medicine, Philadelphia, PA USA; Bioqual Inc., Rockville, MD USA; BioNTech, Mainz, Germany

**Keywords:** Vaccine, mRNA, LNP, COVID-19, SARS-CoV-2, nucleoside-modified, neutralization, Spike, electron micrograph, D614G

## Abstract

The SARS-CoV-2 Spike protein acquired a D614G mutation early in the COVID-19 pandemic that appears to confer on the virus greater infectivity and is now the globally dominant form of the virus. Certain of the current vaccines entering phase 3 trials are based on the original D614 form of Spike with the goal of eliciting protective neutralizing antibodies. To determine whether D614G mediates neutralization-escape that could compromise vaccine efficacy, sera from Spike-immunized mice, nonhuman primates and humans were evaluated for neutralization of pseudoviruses bearing either D614 or G614 Spike on their surface. In all cases, the G614 pseudovirus was moderately more susceptible to neutralization. The G614 pseudovirus also was more susceptible to neutralization by monoclonal antibodies against the receptor binding domain and by convalescent sera from people known to be infected with either the D614 or G614 form of the virus. These results indicate that a gain in infectivity provided by D614G came at the cost of making the virus more vulnerable to neutralizing antibodies, and that the mutation is not expected to be an obstacle for current vaccine development.

## Introduction

There is an urgent worldwide need to develop a safe and effective vaccine for COVID-19. SARS-CoV-2, the etiologic agent of COVID-19, is a novel coronavirus that was first reported in Wuhan, China in December 2019 and four months later was declared a pandemic by the World Health Organization. Vaccine efforts began shortly after the first sequence of the virus was made available in January 2020 (Lurie et al., 2020). Presently, at least 30 SARS-CoV-2 vaccines are in clinical trials (https://www.raps.org/news-and-articles/news-articles/2020/3/covid-19-vaccine-tracker). Because of the urgency and time required to develop a vaccine, most COVID-19 vaccines entering phase 3 trials are based on an early index strain of the virus. These vaccines focus on the viral Spike protein with the goal of eliciting protective neutralizing antibodies (Jackson et al., 2020; Mulligan et al., 2020).

The trimeric Spike protein mediates virus attachment and entry into host cells (Yuan et al., 2020). Each monomer is comprised of an S1 subunit, which contains the receptor binding domain (RBD), and an S2 subunit that mediates membrane fusion (Wrapp et al., 2020). The RBD is the primary target of neutralizing antibodies, though the N-terminal domain and other regions of Spike are also known to possess neutralization epitopes (Brouwer et al., 2020; Chi et al., 2020; Ju et al., 2020b; Liu et al., 2020; Pinto et al., 2020; Shi et al., 2020; Wec et al., 2020; Wu et al., 2020; Yuan et al., 2020).

Early in the pandemic, the virus acquired a D614G mutation in the Spike protein that rapidly increased in frequency and is now the dominant form of the virus globally (Biswas and Majumder, 2020; Gong et al., 2020; Isabel et al., 2020; Islam et al., 2020; Korber et al., 2020a; Koyama et al., 2020a; Koyama et al., 2020b; Mercatelli and Giorgi, 2020). The pattern of spread, combined with increased infectivity in vitro, suggests the mutation gave the virus a fitness advantage for transmission (Korber et al., 2020a; Zhang et al., 2020), despite the fact that this mutation is outside of the RBD in the S1 subunit of Spike. The mutation is also associated with higher virus loads in respiratory secretions but does not appear to increase disease severity (Korber et al., 2020a). A critical question is whether this mutation also mediates neutralization-escape that may reduce the effectiveness of vaccines in general and especially ones based on the D614 version of the Spike immunogen.

Here, we addressed this question using sera from Spike-immunized mice, nonhuman primates (NHP) and humans. Neutralization was measured in 293T/ACE2 cells using lentivirus particles pseudotyped with full-length SARS-CoV-2 Spike containing either D614 or G614. As this was the only difference between the pseudoviruses, any change in phenotype can be directly attributed to D614G. Assays were performed with nearly equivalent amounts of input virus doses (relative light units (RLU) in virus control wells were 501-840x background in all assays).

## Results

Mice, rhesus macaques, and humans were immunized with the nucleoside-modified mRNA-LNP vaccine platform (Alameh et al., 2020; Pardi et al., 2018a). Four different variants of the Spike protein were used as mRNA encoded immunogens; 1) monomeric secreted RBD protein (Laczko et al., 2020), 2) trimeric RBD protein made by adding aT4 fibritin-derived “foldon” trimerization domain to increase immunogenicity (Guthe et al., 2004), 3) prefusion diProline stabilized, first developed with the MERS Spike immunogen (Pallesen et al., 2017), full length cell associated Spike protein (Kirchdoerfer et al., 2016; Kirchdoerfer et al., 2018; Wrapp et al., 2020) and 4) Wuhan index strain (Lurie et al., 2020) with a mutated furin cleavage site cell associated trimeric Spike (Laczko et al., 2020). The furin mutant potentially stabilizes the full-length Spike and maintains the association of the S1 and S2 subunits (Kirchdoerfer et al., 2016). Pseudoviral neutralization titers were calculated as 50% and 80% inhibitory doses (ID50 and ID80, respectively).

A total of forty mice were immunized twice at a 4-week interval with nucleoside-modified mRNA encoding the diProline stabilized cell surface D614 Spike (Kirchdoerfer et al., 2016; Kirchdoerfer et al., 2018; Pallesen et al., 2017; Wrapp et al., 2020) in four equal sized groups, with varying dose and route of vaccine administration. This is the immunogen being employed in the Moderna vaccine in phase 3 clinical trials (mRNA-1273, NCT04470427) (Jackson et al., 2020). Immunizations were performed by the intradermal (I.D.) and intramuscular (I.M.) routes with 10 and 30 µg doses. Preimmune and serum 4 weeks after the second immunization were analyzed for neutralization of pseudoviruses with the D614 and G614 sequences. Preimmune sera from all mice scored negative. At 4 weeks after the second immunization, a relative increase in neutralizing antibody (NAb) titer for G614 over D614 was observed for each animal (Figure 1, Table S1); analyses were done both as a single group and separately by dose and route. Across all routes and doses (N=40), the geometric mean for the G614:D614 ratio was 5.2-fold (95% confidence interval (CI) 4.5 – 6.0) for ID50 and 4.4 (95% CI 4.0-4.9) for ID80. Similar patterns were seen across the 10 µg and 30 µg doses and for both I.M. and I.D. routes. Figure 1 shows the geometric mean for the ratio of G614:D614 NAb titers across route and doses, which ranged from, 3.9 to 6.6 ID50 and 4.3-4.6 ID80. We found no exception to the increased sensitivity of G614 Spike to vaccine sera, even though the D614 form was used for immunization, with p << 0.001 for all comparisons. The maximum percent inhibition was 100% for all animals against both viral strains.

**Figure 1.**
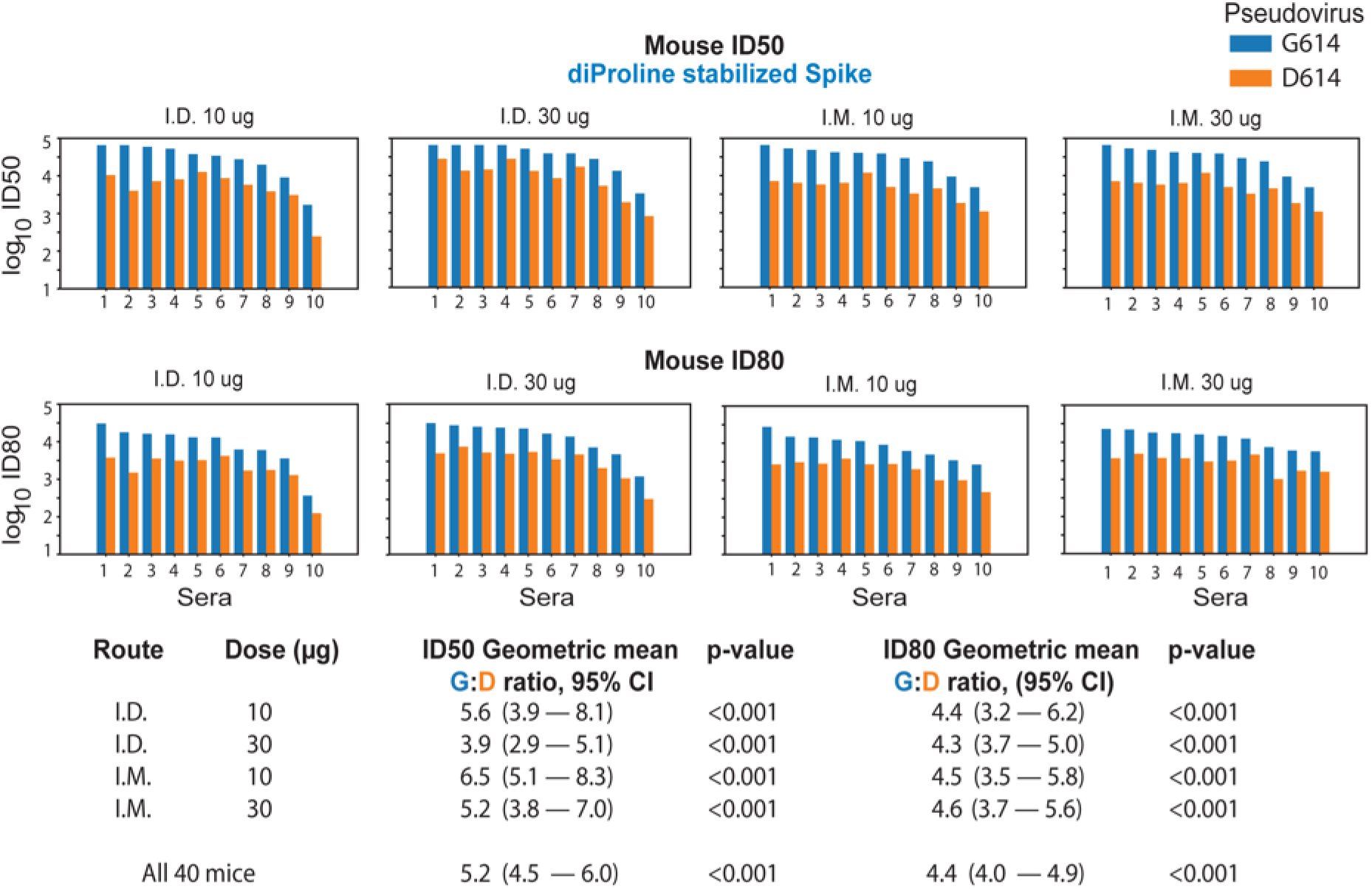
*The G614 Spike is neutralized more potently than the D614 Spike by sera from mice immunized twice at a 4-week interval with nucleoside-modified mRNA-LNPs encoding the Wuhan sequence of Spike (D614) with diProline stabilization mutations. Sera (10 animals/group) obtained 4 weeks after the second immunization were tested for neutralization against pseudoviruses with the D614 and G614 variants of Spike. Log_10_ of the inhibitory dose to reduce infection by 50% (ID50) and by 80% (ID80) are shown; higher values indicate increased neutralization activity. Each pair of bars represents serum from one animal; for each serum the blue bar shows the neutralization titer against the G614 form, the orange bar is the titer against the original D614 form. Top panels are ID50; bottom panels are ID80. Summary statistics for each group are shown at the bottom. The geometric means for the ratio of G614:D614 neutralizing antibodies are shown. Log_10_ of values of the ID50 and ID80 titers were used in a paired t-test to calculate the p-value and the 95% CI of geometric mean for the ratio of G614:D614*.

Eleven rhesus macaques were immunized with the nucleoside-modified mRNA-LNP vaccine platform using 2 different immunogens by the I.M. route (50 µg). The first encoded the Wuhan index strain sequence (Lurie et al., 2020) cell surface Spike protein with a mutated furin cleavage site (furin mut). The second encoded the RBD domain as a secreted monomer. Sera obtained at baseline and 4 weeks after the second immunization were assessed for neutralization of the D614 and G614 variants (Figure 2, Figure S1B and C, Table S1). Baseline (pre-immune) sera were all negative. NAb titers in all post-immunization sera were higher against the G614 variant virus than the D614 (Wuhan index strain) virus for both immunogens. For the furin mut Spike immunogen (N=5), the geometric mean for the G614: D614 ID50 NAb titer ratio was 5.4-fold (95% CI, 3.0 – 9.8) and for ID80 was 2.9-fold (95% CI 2.5-3.3) (Figure 2). For the soluble RBD (N=6), the G614:D614 ID50 NAb titer ratio had a geometric mean of 6.45-fold (p < 0.001) and for ID80 was 3.22-fold (p< 0.001) (Figure 2). Similar to the data in immunized mice, all post-immune NHP sera neutralized both the D614 and G614 viruses to 100%. The observation that the G variant was more sensitive to antibodies induced by both a D614 containing cell surface Spike trimer and an RBD secreted monomer suggests that the G614 mutation increases RBD mediated neutralization despite the fact that D614G lies in S1 outside the RBD.

**Figure 2.**
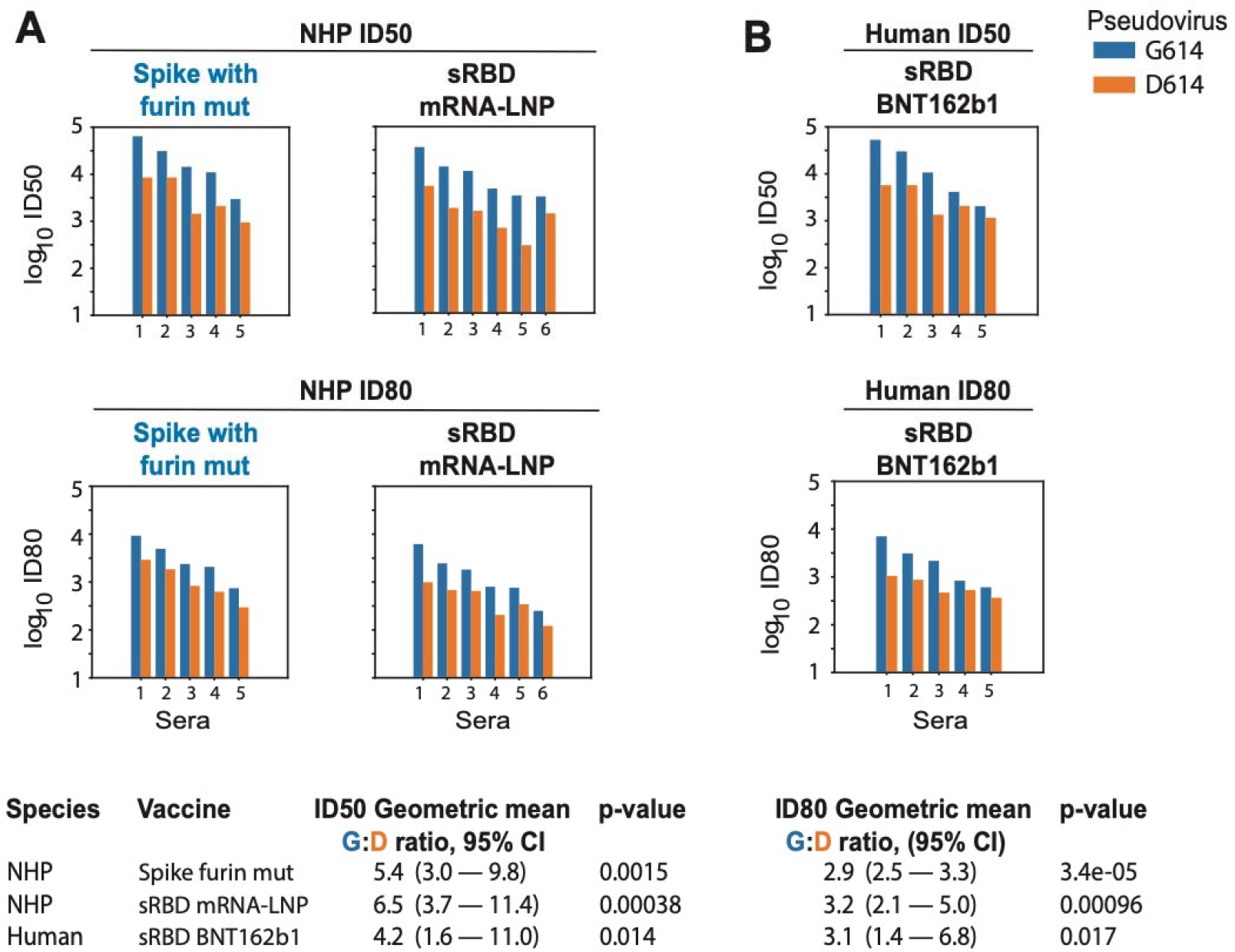
*The G614 Spike is neutralized more potently than the D614 Spike by sera from rhesus macaques (NHP) immunized with nucleoside-modified mRNA-LNPs encoding RBD and full-length Spike immunogens, and humans immunized with RBD trimers. Sera from macaques immunized twice at a 4-week interval with the Wuhan sequence of Spike (D614) with a mutated furin cleavage site (N= 5) or secreted RBD monomers (N= 6) obtained 4 weeks after the second immunization were tested for neutralization against pseudoviruses with the D614 and G614 variants of Spike (A). Sera from 5 humans immunized twice at a 3-week interval with nucleoside-modified mRNA-LNPs encoding a secreted RBD trimer (B). Each pair of bars represents one macaque or human. Top panels are ID50; bottom panels are ID80. For each serum, the blue bar shows the neutralization sensitivity of the G614 form, the orange the original D614 form. The geometric means for the ratio of G614:D614 neutralizing antibody titers measured in sera are provided in the summary at the bottom. Log_10_ of values of the ID50 and ID80 titers were used in a paired t-test to calculate the p-value and the 95% CI of geometric mean for the ratio of G614:D614. Overall response levels were comparable between the two different immunogens in the NHP and between NHPs and humans*.

Preliminary results from a phase 1/2 clinical trial using the nucleoside-modified mRNA-LNP vaccine platform that delivered a secreted RBD trimer were recently published and demonstrated potent ELISA binding and neutralization in all subjects at all tested doses, 10, 30, and 100 μg (Mulligan et al., 2020). Virus neutralizing titers after the second immunization of 10 and 30 μg were 1.8-fold and 2.8-fold greater, respectively, than convalescent sera from SARS-CoV-2 infected patients (Mulligan et al., 2020), although the NAb titers in convalescent patients exhibited a wide range and the patients were older than vaccinated subjects.

Sera obtained preimmunization and 7 days after the second immunization with 50 μg (N=3), 30 μg (N=1), and 10 μg (N=1) of human subjects from a phase 1 trial performed in Germany using the same vaccine were analyzed for neutralization of the D614 and G614 Spike variant pseudoviruses. Similar to the data in murine and NHP immunizations (Figures 1, 2A and S1A-C, Table S1), the G614 pseudovirus was more potently neutralized compared to the D614 pseudovirus by the same serum samples (Figure 2B and S1D, Table S1). The geometric mean of the G614:D614 ID50 NAb titer ratio was 4.22-fold (p = 0.014) and for ID80 was 3.1-fold (p = 0.017). No neutralizing activity was detected in the corresponding preimmune sera. As with the vaccinated mice and NHP, all human vaccine sera achieved complete neutralization of the virus (100% MPI).

We also compared the neutralization susceptibility of D614 and G614 pseudoviruses using convalescent serum samples from people known to be infected with either the D614 or G614 variant of the virus. As shown in Figure 3, the G614 pseudovirus was moderately more sensitive to neutralization by sera from people infected with either variant; this difference was highly statistically significant for ID50, ID80 and maximum percent inhibition (MPI).

**Figure 3.**
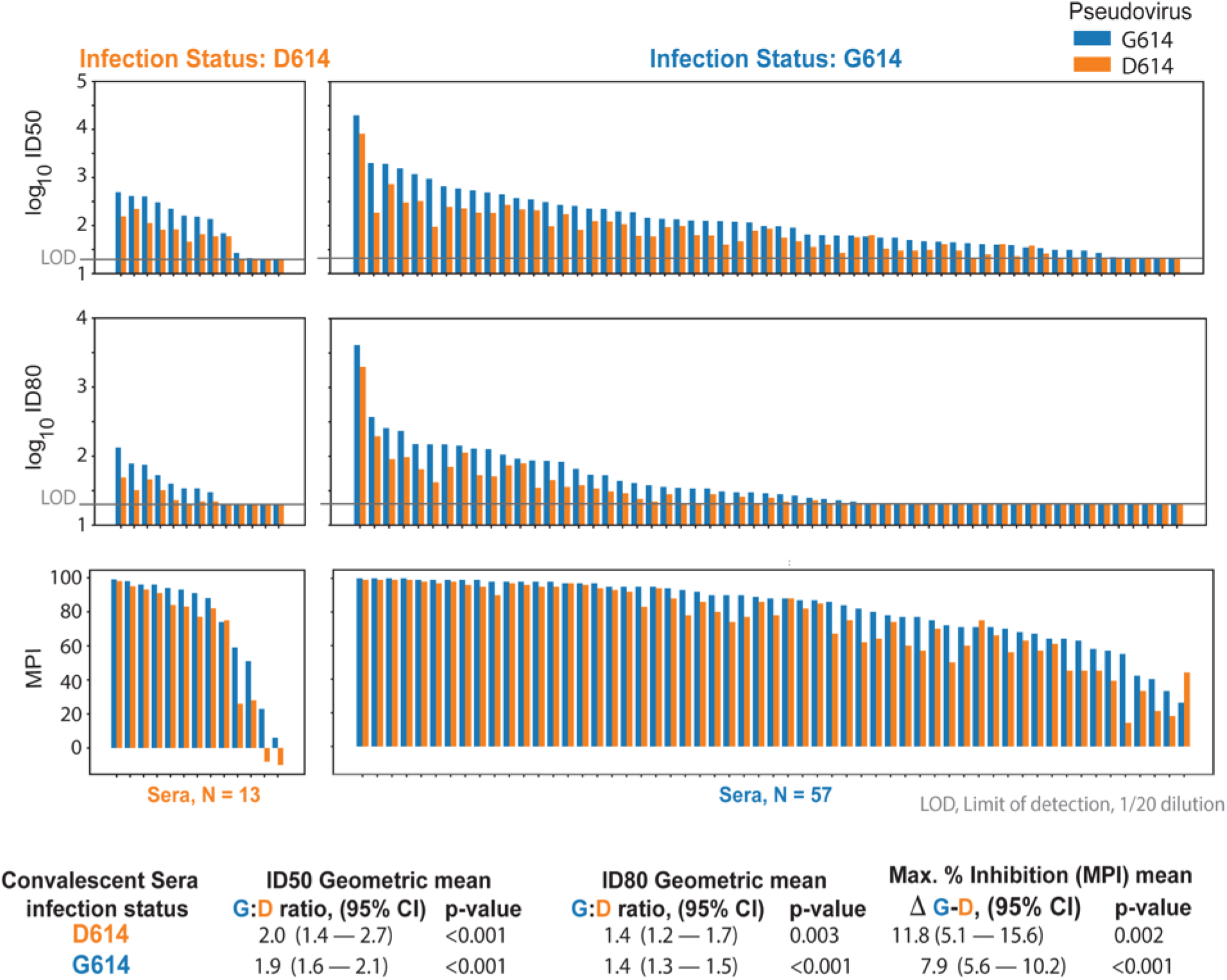
*G614 Spike-pseudotyped virus is neutralized more potently than D614 Spike-pseudotyped virus by sera from people infected with either the D614 or G614 variant of Spike. Each blue/orange pair of bars represents convalescent serum sampled from one person. Left, people infected with D614; Right, people infected with G614. MPI, maximum percent inhibition*.

Additional comparisons of the neutralization susceptibility of D614 and G614 pseudoviruses were made using RBD-specific monoclonal antibodies (mAbs) derived from SARS-CoV-1 and SARS-CoV-2 infected individuals. Antibody CR3022, a weak RBD binder that is reported to be non-neutralizing (ter Meulen et al., 2006) (Yuan et al., 2020), was negative against both pseudoviruses (Figure 4). Antibody B38 was weakly neutralizing against both viruses but 2-fold more potent against G614. Antibodies H4, P2B-2F6 and S309 were 3-142 times more potent against G614. These results are further evidence that G614 is more sensitive to neutralization by RBD-directed antibodies.

**Figure 4:**
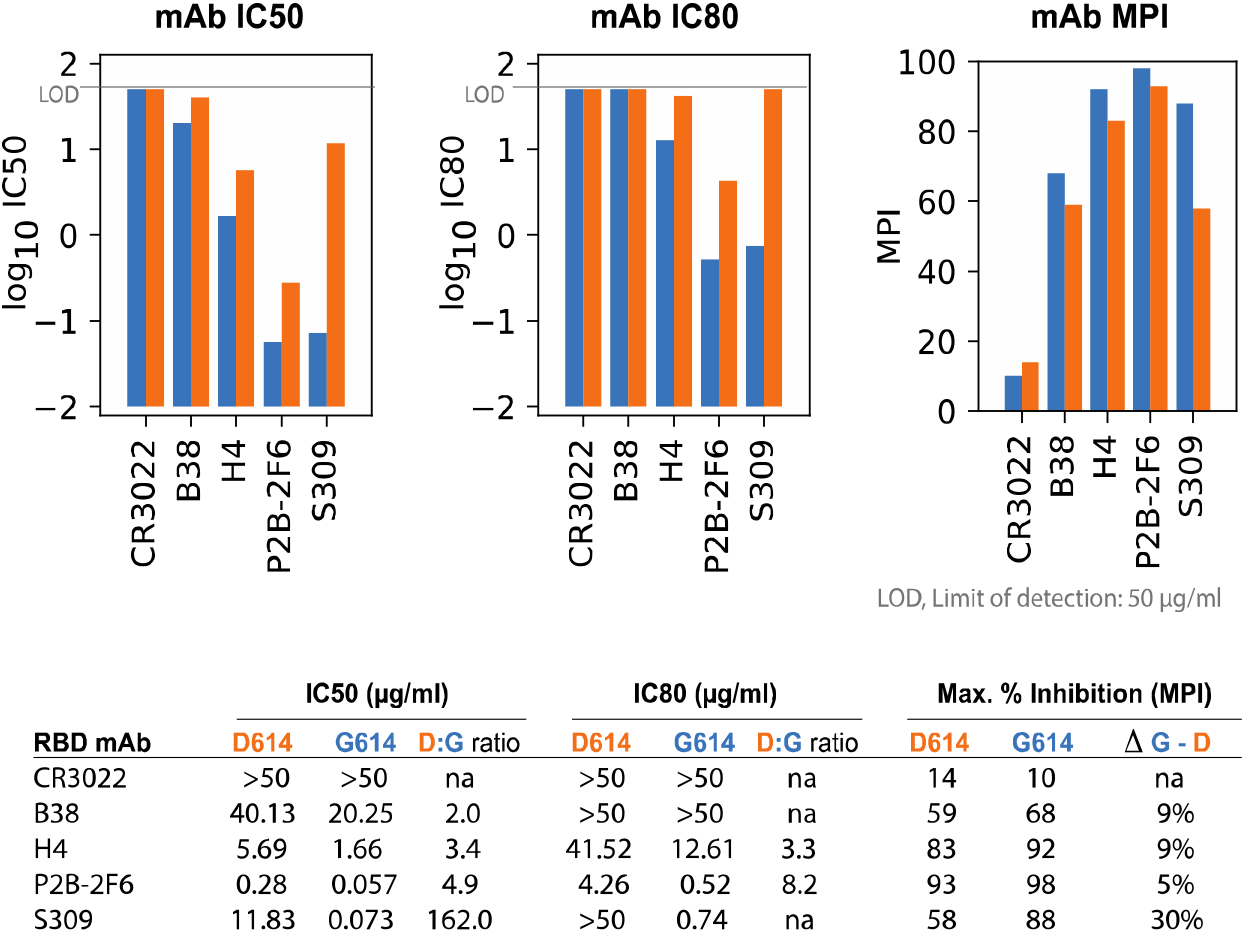
*Differential neutralization of D614 and G614 Spike-pseudotyped viruses by RBD-specific mAbs. MAbs were assayed at 3-fold dilutions starting at 50 µg/ml for a total of 8 dilutions. IC50 and IC80 values are in µg/ml, where a lower bar height corresponds to greater neutralization potency. MPI was calculated as the % neutralization at the highest mAb concentration tested*.

The observation that sera from macaques and humans immunized with RBD-only immunogens more potently neutralized G614 pseudovirus even though the D614G mutation was not in the RBD suggested that the mutation induced a structural change in the expressed Spike that increased the exposure of neutralizing epitopes on the RBD. To evaluate this possibility, we used negative stain electron microscopy (EM) and single particle reconstruction coupled with 3D classification to determine the structure and variability of the two variants in furin-deficient Spike ectodomain constructs (Figure S2). The D614 variant showed two classes of structures called “3-down” or “1-up” in reference to their RBD positions (Figure 5A). The D614 variant demonstrated a roughly equal proportion of the 3-down to 1-up states (54% versus 46%), consistent with cryo-EM data reported by others (Walls et al., 2020). The G614 variant also demonstrated 3-down and 1-up structures (Figure 5B); however, the 1-up state was more heavily populated, 82%. This shift to the 1-up state as the dominant form in the Spike ectodomain demonstrates an allosteric effect of the D614G mutation on the RBD conformation and suggests a mechanism for the enhanced neutralization susceptibility of the G614 variant.

**Figure 5.**
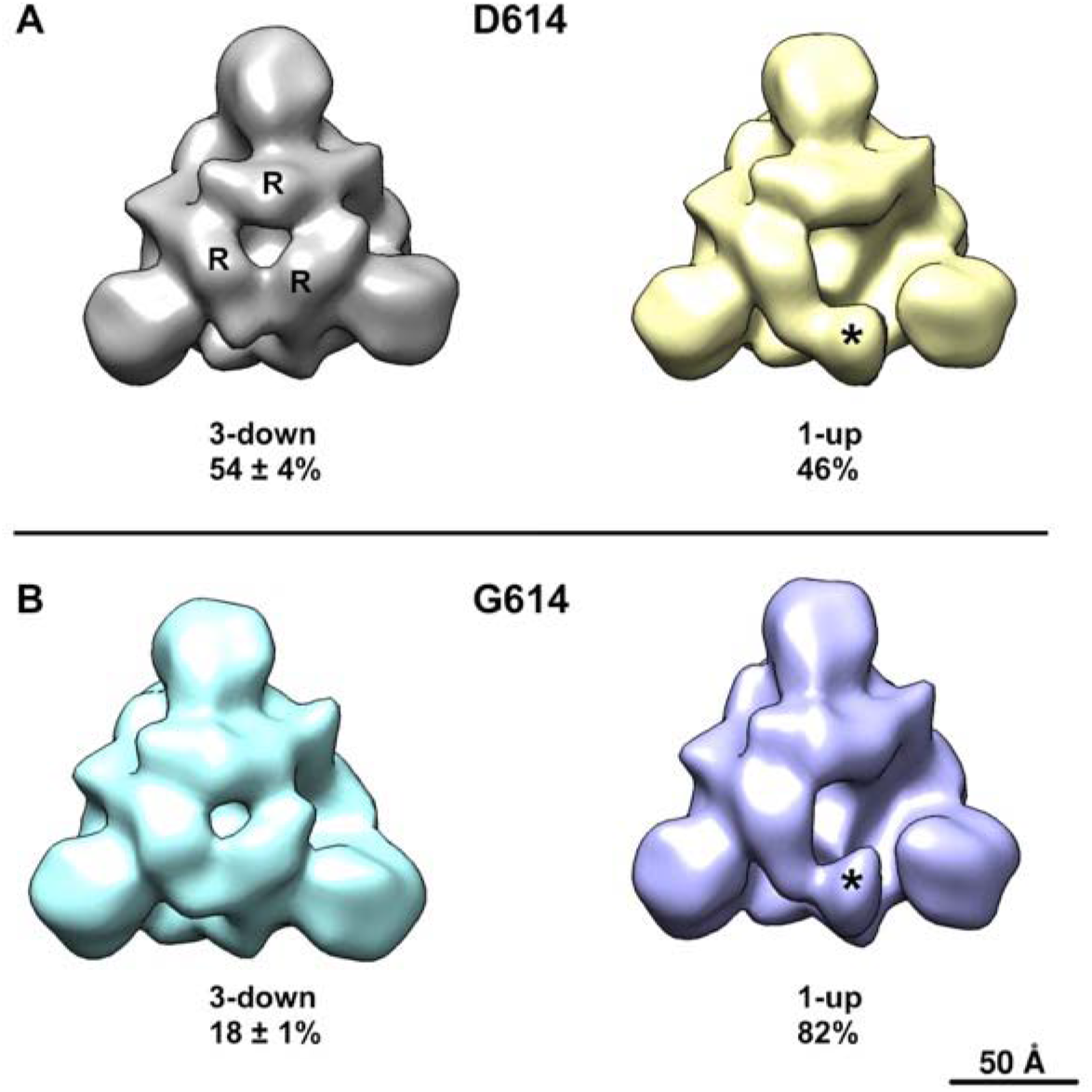
*Negative stain electron microscopy reconstructions of expressed Spike constructs following 3D classification. View is looking down the 3-fold trimer axis onto the S1 domain. (A) D614 variant showing the 3-RBD-down structure on the left with individual RBDs labeled (R), and the 1-RBD-up structure on the right with the up RBD labeled (asterisk). Fraction of particle images that sorted into each class indicated below, expressed as average* ± *standard deviation, N=3 each. (B) G614 variant also showing 3-down and 1-up structures*.

The preferred occupancy of the 1-up state of the G614 variant raised the possibility that G614 infections might elicit more potent antibody responses as a consequence of increased exposure of RDB epitopes during infection. Given the complexity of the data and the potential for antibody responses to decline over time in convalescent sera (Ibarrondo et al., 2020), we explored this possibility using a mixed effect model to predict log ID50. We modeled the different levels of responses in sera from different individuals (N=70) as a random effect and used fixed effects to model the contribution the number of days between diagnosis and sera sampling, the RT PCR cycle threshold (Ct) values, the pseudovirus tested, and the D and G infecting strain. While the relationship between the pseudovirus tested and the ID50 was highly significant (Sup. Figure S3), consistent with the paired statistic in Figure 3, we found that none of the other factors, including interactions between the D or G infecting virus and pseudovirus assayed were statistically significant predictors.

## Discussion

Soon after the G614 mutation in Spike appeared early in the pandemic, it rapidly replaced the D614 variant in many countries (Korber et al., 2020a). This mutation is associated with increased infectivity (Korber et al., 2020a; Zhang et al., 2020) when tested in in vitro model systems. Over 100 vaccines using various platforms and immunogens are being developed to combat COVID-19 and end the devastating financial, societal, and health burdens. Currently, over 30 vaccines are in clinical testing, some of which have entered phase 3 trials. Most SARS-CoV-2 vaccines were originally designed using the D614 variant of the Spike protein, which was present in the first sequence of SARS-CoV-2 from Wuhan (Lurie et al., 2020). The most critical finding that will ease the concern for most current vaccines in clinical trials is our data showing that the SARS-CoV-2 Spike protein with the G614 mutation does not escape neutralization but rather is neutralized at a higher level by serum from vaccinated mice, NHPs, and humans that used immunogens derived from the D614 variant of the virus. Consistent with this finding, our data show that the G614 variant of the virus was more sensitive to neutralization by serum samples from people known to be infected with either variant.

D614 is on the surface of the Spike protomer and has the potential to influence the conformation and flexibility of the Spike protein. The recently published cryo-EM structure of the SARS-CoV-2 Spike demonstrates that the D614 sidechain can form a hydrogen bond with the neighboring protomer T859 amino acid (Wrapp et al., 2020). This interaction may be critical, as it may bridge residues from the S1 region of one protomer to the S2 region of an adjacent protomer. This interaction would bracket the furin and S2 cleavage sites. Potentially, it could reduce shedding of S1 from viral-membrane-bound S2 and the introduction of G614 could increase S1 release. Our structural data demonstrate that, in the context of the stabilized ectodomain, this mutation leads to an increase in the percent of 1 RBD region per trimer being in the up position (Figure 5), which is necessary for binding to ACE2 and infection of target cells. A recent publication demonstrated a similar effect of the G614 mutation to increase the number of RBDs in the up position (Yurkovetskiy et al., 2020). Using an alternative structural analysis method, extensive microsecond timescale atomistic molecular dynamics simulations, reveal that in the G-form, the inter-protomer interactions in the Spike trimer become more symmetric compared to the D-form. This equalization of inter-protomer energetics results in a higher population of one-up Spike conformations, leading to increased encounter between RBD and ACE2 receptor and greater exposure of RBD domain for neutralization (Mansbach et al., 2020).

Our results in immunized mice, NHPs and humans subjects immunized with nucleoside-modified mRNA-LNP vaccines with various Spike immunogens, humans known to be infected with either the D614 or G614 variant of the virus, and with RBD-specific monoclonal antibodies conclusively demonstrate a modest but highly consistent increase in neutralization-susceptibility of the G614 variant. A clinical trial (Sahin et al., 2020) and a preclinical analysis (Corbett et al., 2020) also using nucleoside-modified mRNA-LNP COVID-19 vaccines have been published after submission of this manuscript. They both analyzed neutralization of D614 and G614 viruses with assays that utilized pseudotyped Vesicular Stomatitis Virus. Both observed no statistical reduction in neutralization of the G614 variant compared to D614, but did not directly compare neutralization of each virus by serum from individual subjects or animals. Assays performed in Erica Ollmann-Saphire’s laboratory demonstrate equivalent or better neutralization of G614-bearing pseudovirus compared to D614-bearing pseudovirus using convalescent sera from 6 COVID-19 subjects, but it was not known whether the individuals were infected with the D614 or G614 variant (Korber et al., 2020b).

A conclusion from our data is that the virus evolved to be more easily neutralized by host antibody responses. Previous work has shown that the G614 virus variant is more infectious in vitro (Korber et al., 2020a; Zhang et al., 2020); given this, the fitness advantage for the virus due to the D614G mutation is related to enhanced infectivity, in the setting of increased neutralization sensitivity. Given that the mutation is in Spike, it was important to determine if the newer globally dominant form of the virus was antigenically different as well; a priori, one would assume that the mutation might be more resistant to neutralization. What we found instead is that the new form of Spike is actually *more* sensitive to antibody neutralization by vaccine and infection induced immune responses. The fact that the mutation is outside of the RBD and fusion machinery and affects both infectivity and neutralization is common for viral fusion proteins. Thus, coronavirus Spike proteins are prefusion stabilized by similar types of mutations. Also, a variety of mutations distinct from neutralization epitopes in HIV envelope are known to alter antigenicity (Review in Medina-Ramirez et al., 2017).

Potential drawbacks to our studies include: 1) while we studied 4 different variations of the Spike immunogen, we only used a single type of vaccine platform, nucleoside-modified mRNA-LNP platform (reviewed in (Alameh et al., 2020; Pardi et al., 2018b)). This platform is recognized as an outstanding new approach that has entered phase 3 clinical trials by 2 separate pharmaceutical/biotech companies, Moderna and Pfizer/BioNTech. The results of phase 1 clinical trials demonstrated that all immunized subjects safely developed neutralizing responses (Jackson et al., 2020; Mulligan et al., 2020). 2) We performed pseudovirus neutralization assays. While these assays are considered excellent methods to measure neutralization and are used for the development of many viral vaccines, live virus neutralization or animal challenges would offer additional methods to measure effective vaccine response to the G614 variant. A new study directly compared 2 different pseudovirus neutralization assays, including lentiviral used in this study, with live virus neutralization (Schmidt et al., 2020). Highly significant correlations were observed between the assays demonstrating no significant differences in the use of live virus versus pseudovirus neutralization assays for both convalescent plasma and human monoclonal antibodies, which is perhaps not surprising given that neutralization in both assays is based on entry inhibition. As with the pseudovirus assay, assays with live SARS-CoV-2 are performed with viruses that are produced and assayed in cell lines; neutralization assays that use natural target cells of the respiratory system are technically challenging, low throughput and difficult to standardize. 3) Our structural studies were performed in the context of a furin cleavage-deficient Spike ectodomain. While this soluble ectodomain has been shown to be a good mimic of the native Spike, and the shift in the proportion of RBD “up” conformation between the D614 and G614 variants suggest an allosteric effect of the D614G mutation on the RBD conformations, the structures of the native Spike may have some differences from what we observe in the context of the ectodomain.

In summary, we demonstrate here that vaccinated mice, NHPs, and humans using the nucleoside-modified mRNA-LNP vaccine platform encoding 4 different SARS-CoV-2 Spike immunogens generate antibody responses that not only recognize the G614 mutation that has taken over the pandemic (Zhang et al., 2020)(Korber et al., 2020a), they have stronger titers of neutralization to this virus variant. The mechanism appears to be that the mutation increases the up formation of the RBD in the Spike trimer, increasing the exposure of neutralization epitopes. Over thirty vaccines are currently in clinical trial testing. Most of the immunogens in these trials were either derived from the initial D614 virus or contain D614G in the Spike. While the G614 variant has replaced the original D614 sequence in the SARS-CoV-2 Spike throughout much of the world, our finding that this is not an escape mutation and, in fact, is better neutralized by sera from mice, NHPs, and humans immunized with vaccines derived from the D614 viral Spike alleviates a major concern regarding the current efforts to develop an effective SARS CoV-2 vaccine.

## Data Availability

I will share all primary data generated as part of this grant. It will be deposited at the National Addiction & HIV Data Archive Program (NAHDAP), which is an NIH-funded repository. Collected samples will be stored at the University of Pennsylvania and available after review of the request by the PI.

https://www.icpsr.umich.edu/icpsrweb/NAHDAP/data/index.jsp

## AUTHOR CONTRIBUTIONS

We would like to acknowledge members of the Sheffield COVID-19 Genomics Group for help during SARS-CoV-2 sequencing, and the COVID-HERO research team for help with participant recruitment and sample processing. We thank Peter Kwong (Vaccine Research Center, NIH) for providing the monoclonal antibodies. We also thank James Theiler for statistical advice.

## AUTHOR CONTRIBUTIONS

M-G. A., C. C. LaB., R. J. E., L. S., S. S., K. M., S. G., C. M., N. P. conducted the experiments, D.W., T. deS., P.C., H.H., R.B., N. H., P. J. C. L., Y. T., P. A. S., M-G. A., M. G. L., P. A., B. F. H., B. K., D. C. M. designed the experiments, C.B.; U.S. performed the phase 1 clinical trial, T. deS., P.C., H.H., R.B. obtained and supplied samples from infected patients and D.W., B. K., P. A. S., P. A., D. C. M. wrote the paper, and all authors reviewed and edited the manuscript.

## START METHODS

### Key Resources Table

| REAGENTS or RESSOURCE | SOURCE | IDENTIFIER |
| --- | --- | --- |
| **Antibodies** |  |  |
| CR3022 | Peter Kwong, VRC/NIH |  |
| B38 |  |  |
| H4 |  |  |
| P2B-2F6 |  |  |
| S309 |  |  |
| **Chemicals and Recombinant Proteins** |  |  |
| M1-Pseudouridine 5'-triphosphate | TriLink | N-1081 |
| CleanCap | TriLink | N-7413 |
| Cellulose | Sigma-Aldrich | 11363-250G |
| MEGAscript™ T7 Transcription Kit | ThermoFisher Scientific | AMB13345 |
| QuikChange Lightning Site-Directed Mutagenesis Kit | Agilent | 210518 |
| **Experimental Models: Organisms/strains** |  |  |
| BALB/c mouse | Charles Rivers Laboratories | N/A |
| **Experimental Models: Cell Lines** |  |  |
| 293T/ACE2 cells | Drs. Mike Farzan and Huihui Mu at Scripps | 293T/ACE2 cells |
| **Bacterial and Virus strains** |  |  |
| **Recombinant DNA** |  |  |
| pTEV-di-proline SARS-CoV-2 Spike-A101 | Weissman Lab |  |
| pTEV full length S-furin mutant-A101 | Weissman Lab |  |
| pTEV-RBD-A101 | Weissman Lab |  |
| **Software and Algorithms** |  |  |

## RESOURCE AVAILABILITY

### Lead Contact

Further information and requests for supporting data, resources, and reagents should be directed to and will be fulfilled upon request by the Lead Contacts: Drew Weissman or David Montefiori

### Materials Availability

Reagents from this study are available upon request.

### Data and Code Availability

Data from this study are available upon request.

## EXPERIMENTAL MODELS AND SUBJECT DETAILS

### Mice

BALB/c mice aged 8 weeks old were purchased from Charles Rivers Laboratories and immunized after 1 week of acclimation at the University of Pennsylvania.

### Nonhuman Primate

NHP studies were performed at Bioqual, Inc, Rockville, MD following their and University of Pennsylvania’s IACUC approval.

### Human

Serum from subjects enrolled in a phase 1/2 clinical trial of a nucleoside-modified mRNA-LNP vaccine encoding trimeric SARS-CoV-2 RBDs (BNT162b1)(Mulligan et al., 2020) were obtained (NCT04368728).

Serum samples from people known to be infected with either the D614 or G614 form of SARS-CoV-2 were collected following informed consent from healthcare workers at Sheffield Teaching Hospitals NHS Foundation Trust, Sheffield, UK as part of the COVID-HERO SARS-CoV-2 seroprevalence study (Research Ethics Committee reference 20/HRA/2180).

### Cell lines

293T/ACE2 cells were kindly provided by Drs. Mike Farzan and Huihui Mu at Scripps.

## METHOD DETAILS

### Ethics statement

The clinical trial described in this manuscript was randomized double-blinded and the sera collections were approved by the appropriate IRBs. All protocols, experimentation and animal manipulation adhered to the Guide for the Care and Use of Laboratory Animals by the Committee on Care of Laboratory Animal Resources Commission on Life Sciences, National Research Council under IACUC approved protocols.

### mRNA production

N^1^-methylpseudouridine modified mRNA was produced as previously described (Freyn et al., 2020) using T7 RNA polymerase (MegaScript, ThermoFisher Scientific, Waltham, MA, USA) on Not-I/AlfII double digested and linearized plasmids encoding codon-optimized di-proline modified pre-fusion SARS-CoV-2 Spike (Wuhan Hu-1 complete genome, GenBank: MN908947.3), full length S-furin mutant protein (RRAR furin cleavage site abolished), and RBD. In vitro transcribed (IVT) mRNAs were co-transcriptionally capped using the CleanCap system (TriLink Biotechnologies, San Diego, CA, USA), and purified using a cellulose-based chromatography method (Baiersdorfer et al., 2019). All IVT mRNAs were analyzed on agarose (1.4 % w/v, 1X TAE buffer) for integrity, and subjected to additional quality control to rule out double stranded RNA (dsRNA) contamination and endotoxin contamination prior to formulations into lipid nanoparticles (LNPs), as described (Pardi et al., 2015).

### Production of mRNA-LNP

Lipid nanoparticles (LNP) used in this study contain an ionizable lipid proprietary to Acuitas /DSPC/Cholesterol/PEG-Lipid (Lin et al., 2013). Encoding mRNA was encapsulated in LNP using a self-assembly process in which an aqueous solution of mRNA at 4.0 pH was rapidly mixed with a solution containing the aforementioned lipids premixed and dissolved in ethanol. The proprietary lipid and LNP composition are described in US patent US10,221,127. All LNP were characterized post-production at Acuitas pharmaceutical (Vancouver, BC, Canada) for their size, polydispersity using a Malvern Zetasizer (Zetasizer Nano DS, Malvern, UK), encapsulation efficiency, and shipped on dry ice and stored at −80°C until use.

### Administration of test articles (immunization) and blood collection

8-12 week old Balb/c mice were immunized with either 10 or 30 µg of mRNA-LNP via the intramuscular (I.M.) or intradermal (I.D.) routes of administration using a 29G X1/2’’ Insulin syringe. Mice received a booster injection on day 28 (4 weeks). Blood was collected at day 0, 28 and 56 through the retro-orbital route using non heparinized micro hematocrit capillary tubes (ThermoFisher Scientific, Waltham, MA, USA). Serum was separated by centrifugation (10 000 g, 5 min) using a non-refrigerated Eppendorf 5424 centrifuge (Eppendorf, Enfield, CT, USA), heat-inactivated (56°C) for 30 minutes, and stored at −20° C until analysis.

### Administration of mRNA/LNPs to rhesus macaques

Fifty micrograms of either mRNA-LNP encoding an unstabilized transmembrane (TM) Spike protein with a mutant furin cleavage site or a monomeric soluble RBD were administered I.M. in two sites in the left and right quadriceps on weeks 0, and 4. Animals were anesthetized with ketamine prior to blood draws from the femoral vein. Serum samples were analyzed in 5 macaques immunized with mRNA-LNP encoding unstabilized TM Spike with a mutant furin cleavage site and in 6 animals immunized with mRNA-LNP encoding soluble RBD. All studies were performed at Bioqual, Inc, Rockville, MD following IACUC approval.

### Clinical trial samples

Serum from subjects enrolled in a phase 1/2 clinical trial of a nucleoside-modified mRNA-LNP vaccine encoding trimeric SARS-CoV-2 RBDs (BNT162b1)(Mulligan et al., 2020) were obtained (NCT04368728). Five subjects that received 2 immunizations at a 3-week interval with 10, 30, or 50 µg of mRNA were used. All subjects were considered a single group, as similar serologic data was obtained for each dose and the comparison and calculation of statistical significance was performed within each sample. Serum was obtained prior to the first immunization and 7 days after the second immunization.

Serum samples from people known to be infected with either the D614 or G614 form of SARS-CoV-2 were collected following informed consent from healthcare workers at Sheffield Teaching Hospitals NHS Foundation Trust, Sheffield, UK as part of the COVID-HERO SARS-CoV-2 seroprevalence study (Research Ethics Committee reference 20/HRA/2180). Details of previous RT-PCR confirmed diagnosis of SARS-CoV-2 were recorded. SARS-CoV-2 sequences were generated using samples collected for routine clinical diagnostic use in individuals presenting with COVID-19 at Sheffield Teaching Hospitals NHS Foundation Trust, Sheffield, UK. This work was performed under approval by the Public Health England Research Ethics and Governance Group for the COVID-19 Genomic UK consortium (R&D NR0195). Extracted nucleic acid from nasal or throat swabs was used for whole genome sequencing of SARS-CoV-2 (Oxford Nanopore Technologies (ONT), Oxford, UK) using the ARTIC network protocol (https://artic.network/ncov-2019). Following base calling, data were demultiplexed using ONT Guppy using a high accuracy model. Reads were filtered based on quality and length (400 to 700bp) and mapped to the Wuhan reference genome (MN908947). Variants, including the A-to-G change at nucleotide 23,403 leading to D614G, were called using nanopolish (https://github.com/jts/nanopolish) and consensus sequences compared to the reference.

### Monoclonal antibodies

SARS-CoV-2 RBD-binding mAbs CR3022, B38, H4, P2B-2F6, and S309 were obtained from Dr. Peter Kwong at the Vaccine Research Center, National Institutes of Health, USA. CR3022 was isolated from a convalescent SARS-CoV patient, exhibits cross-reactive binding to SARS-CoV-2 RBD distal from the receptor binding motif, and does not neutralize SARS-CoV-2 even at concentrations as high as 400 µg/ml (ter Meulen et al., 2006; Yuan et al., 2020). B38 and H4 were isolated from a COVID-19 subject, bind different epitopes on RBD that partially overlap, and exhibit potent SARS-COV-2 neutralizing and ACE2-blocking activity (Wu et al., 2020). P2B-2F6 was isolated from a COVID-19 subject in Shenzhen, China, exhibits potent SARS-CoV-2 neutralizing activity and blocks ACE2 binding (Ju et al., 2020a). S309 was isolated from a recovered SARS-CoV-infected subject, potently cross-neutralizes SARS-CoV and SARS-CoV-2, binds outside the receptor binding motif of RBD and, based on cryo-EM structure, is not predicted to interfere with ACE2 binding (Pinto et al., 2020).

### SARS-CoV-2 pseudovirus neutralization assay

SARS-CoV-2 neutralization was assessed with Spike-pseudotyped viruses in 293T/ACE2 cells as a function of reductions in luciferase (Luc) reporter activity. 293T/ACE2 cells were kindly provided by Drs. Mike Farzan and Huihui Mu at Scripps. Cells were maintained in DMEM containing 10% FBS and 3 µg/ml puromycin. An expression plasmid encoding codon-optimized full-length Spike of the Wuhan-1 strain (VRC7480), was provided by Drs. Barney Graham and Kizzmekia Corbett at the Vaccine Research Center, National Institutes of Health (USA). The D614G amino acid change was introduced into VRC7480 by site-directed mutagenesis using the QuikChange Lightning Site-Directed Mutagenesis Kit from Agilent Technologies (Catalog # 210518). The mutation was confirmed by full-length Spike gene sequencing. Pseudovirions were produced in HEK 293T/17 cells (ATCC cat. no. CRL-11268) by transfection using Fugene 6 (Promega Cat#E2692) and a combination of Spike plasmid, lentiviral backbone plasmid (pCMV ΔR8.2) and firefly Luc reporter gene plasmid (pHR’ CMV Luc)(Naldini et al., 1996) in a 1:17:17 ratio. Transfections were allowed to proceed for 16-20 hours at 37°C. Medium was removed, monolayers rinsed with growth medium, and 15 ml of fresh growth medium added. Pseudovirus-containing culture medium was collected after an additional 2 days of incubation and was clarified of cells by low-speed centrifugation and 0.45 μm micron filtration and stored in aliquots at −80°C. TCID_50_ assays were performed on thawed aliquots to determine the infectious dose for neutralization assays (RLU 500-1000x background, background usually averages 50-100 RLU).

For neutralization, a pre-titrated dose of virus was incubated with 8 serial 3-fold or 5-fold dilutions of serum samples or mAbs in duplicate in a total volume of 150 μl for 1 hr at 37°C in 96-well flat-bottom poly-L-lysine-coated culture plates (Corning Biocoat).

Cells were suspended using TrypLE express enzyme solution (Thermo Fisher Scientific) and immediately added to all wells (10,000 cells in 100 μL of growth medium per well). One set of 8 control wells received cells + virus (virus control) and another set of 8 wells received cells only (background control). After 66-72 hrs of incubation, medium was removed by gentle aspiration and 30 μL of Promega 1X lysis buffer was added to all wells. After a 10 minute incubation at room temperature, 100 μl of Bright-Glo luciferase reagent was added to all wells. After 1-2 minutes, 110 μl of the cell lysate was transferred to a black/white plate (Perkin-Elmer). Luminescence was measured using a PerkinElmer Life Sciences, Model Victor2 luminometer. Neutralization titers are the serum dilution (ID50/ID80) or mAb concentration (IC50/IC80) at which relative luminescence units (RLU) were reduced by 50% and 80% compared to virus control wells after subtraction of background RLUs. Maximum percent inhibition (MPI) is the % neutralization at the lowest serum dilution or highest mAb concentration tested. Serum samples were heat-inactivated for 30 minutes at 56°C prior to assay.

### Protein expression and purification

SARS-CoV-2 ectodomain constructs (Wrapp et al., 2020) were produced and purified as follows. Genes encoding residues 1–1208 of the SARS-CoV-2 S (GenBank: MN908947) with a “GSAS” substitution at the furin cleavage site (residues 682–685), with and without proline substitutions of residue K986 and V987 (S-GSAS/PP or S-GSAS), a C-terminal T4 fibritin trimerization motif, an HRV3C protease cleavage site, a TwinStrepTag and an 8XHisTag were synthesized and cloned into the mammalian expression vector pαH. The S-GSAS template was used to include the D614G mutation (S-GSAS(D614G)). Plasmids were transiently transfected into FreeStyle-293F cells using Turbo293 (SpeedBiosystems). Protein was purified on the sixth day post-transfection from filtered supernatant using StrepTactin resin (IBA), followed by size-exclusion chromatography (SEC) purification using a Superose 6 10/300 GL column (GE healthcare) equilibrated in 2mM Tris, pH 8.0, 200 mM NaCl, 0.02% sodium azide buffer.

### Negative-stain electron microscopy

Samples of S-GSAS and S-GSAS (D614G) ectodomain constructs were diluted to 100 µg/ml with room-temperature buffer containing 20 mM HEPES pH 7.4, 150 mM NaCl, 5% glycerol and 7.5 mM glutaraldehyde, and incubated 5 min; then glutaraldehyde was quenched for 5 min by addition of 1M Tris stock to a final concentration of 75 mM. A 5-μl drop of sample was applied to a glow-discharged, carbon-coated grid for 10-15 s, blotted, stained with 2% uranyl formate, blotted and air-dried. Images were obtained with a Philips EM420 electron microscope at 120 kV, 82,000× magnification, and a 4.02 Å pixel size. The RELION program (Scheres, 2016) was used for particle picking, 3D classification, and 3D refinements. The number of particle images that sorted into each class during the classification provides an estimate of the fraction for each state. We have previously shown this method has adequate resolution to resolve and compare up and down conformations (Henderson et al., 2020).

### Statistical methods

For a given experiment, neutralizing antibody titers for G614 and D614 were measured from serum on each animal and compared using a paired t-test of the logarithm of the antibody titers. This formally tests whether the ratio of the titer is different from 1. All statistical tests were performed at the 0.05 level in the R software (v3.6.1) (Team, 2019).

To explore whether the infecting form of the virus impacted the serum potency, we modeled the logarithm of ID50 as a mixed effect model, using a random effect to account for the different levels of responses in sera from different individuals and (Table S2) effects to model the contribution the number of days between diagnosis and sera sampling, the RT PCR cycle threshold (Ct) values, the pseudovirus tested and the D and G infecting strain. Observations below the Limit Of Detection (LOD) were replaced by LOD/V2. The resulting model only includes the effect of the pseudo-strain at a much higher level of confidence (see fixed-effect ANOVA that tests the statistical significance of the pseudo-strain effect). In this analysis, neither viral load (as measured by CT), days between infection and testing, nor interactions between the infecting virus strain and the pseudovirus were statistically significant predictors for log ID50

## Supplementary tables

### Tables in Excel File

**Table S1**. ID50, ID80 and maximum percent inhibition (MPI) values for all serum samples assayed in this study.

**Table S2.** ANOVA table for fixed effects in the fitted mixed effect model. Only the effect of the pseudo-strain is statistically significant. In particular, the interaction between the strain and the pseudo strain is not significant.

|  | Sum Sq | Mean Sq | NumDF | DenDF | F value | P[ > F] |
| --- | --- | --- | --- | --- | --- | --- |
| **Strain D vs G** | 0.0076 | 0.0076 | 1 | 66 | 0.05 | 0.82 |
| **Pseudovirus** | 10.77 | 10.77 | 1 | 68 | 70.54 | 4.1 e-12 |
| **CT value** | 0.052 | 0.052 | 1 | 66 | 0.34 | 0.56 |
| **Diagnosis delay** | 0.157 | 0.157 | 1 | 66 | 1.03 | 0.31 |
| **Strain:Pseudo** | 0.0056 | 0.0056 | 1 | 68 | 0.07 | 0.85 |

## Supplementary figures

**Figure S1.**
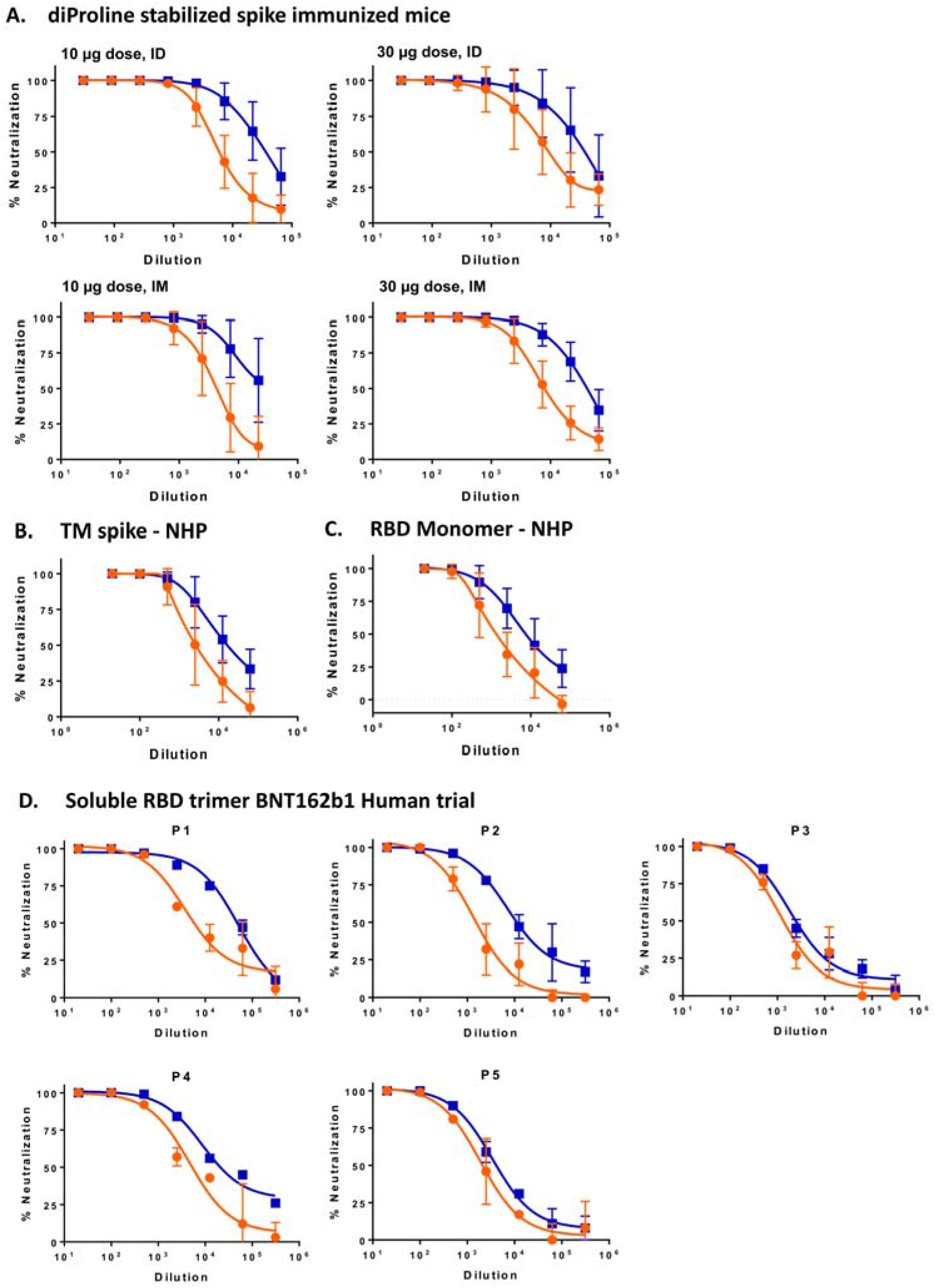
The G614 Spike is more vulnerable to neutralization than D614 Spike by vaccine-elicited serum antibodies. This figure accompanies Figures 1 and 2 and their legends.. **(A, B and C)** Percent neutralization values at each serum dilution for all animals in each group were averaged. **(D)** Individual neutralization curves for the 5 vaccine recipients in a phase 1/2 clinical trial of nucleoside-modified mRNA-LNPs encoding a secreted RBD trimer. Orange, pseudovuirus bearing D614 Spike; Blue, pseudovirus bearing G614 Spike.

**Figure S2.**
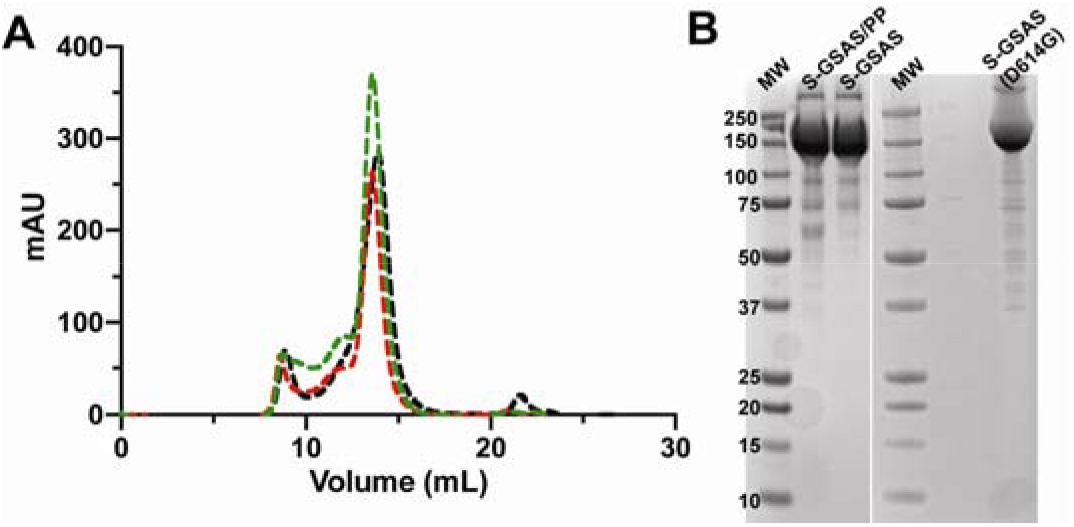
Purification the SARS-CoV-2 S ectodomain. **(A)** Chromatogram of the size-exclusion purification on a Supersose 6 GL 10/300 column of the Spike ectodomain with proline mutations of residues 986 and 987 and furin cleavage site (residues 682-685) mutated to GSAS (S-GSAS/PP; in black), of the S ectodomain with the proline mutations reverted to the native K and V residues (S-GSAS; in red) and the Spike ectodomain with the mutation D614G (S-GSAS(D614G); in green). **(B)** SDS-PAGE of the purified protein (MW: molecular weight marker; 15 µg/lane).

**Figure S3.**
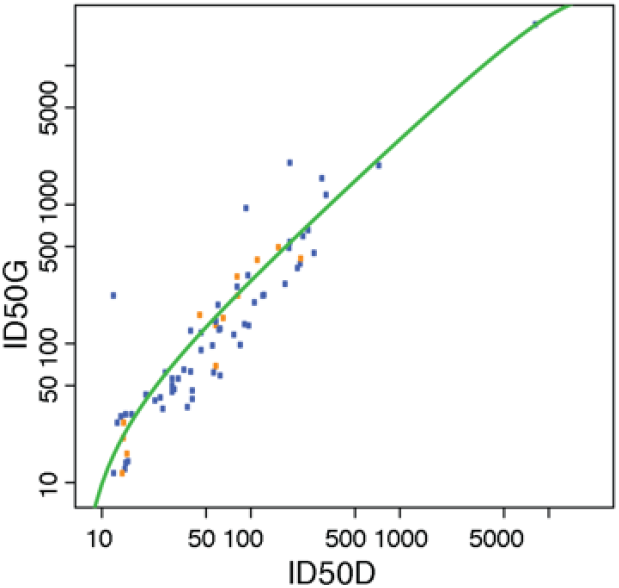
Data for linear model to test whether the infecting strain impacted the potency of the response. Data showing the increased potency of sera against G614 pseduoviruses (y-axis) relative to D614 pseudoviruses (x-axis), and lack of association of overall potency with the infecting variant. The green line is a quadratic fit. The blue dots are from G614 infected individuals and the orange dots from D614 infected individuals.

